# The NYCKidSeq project: study protocol for a randomized controlled trial incorporating genomics into the clinical care of diverse New York City children

**DOI:** 10.1101/2020.09.02.20186361

**Authors:** Jacqueline A. Odgis, Katie M. Gallagher, Sabrina A. Suckiel, Katherine E. Donohue, Michelle A. Ramos, Nicole R. Kelly, Gabrielle Bertier, Christina Blackburn, Kaitlyn Brown, Lena Fielding, Jessenia Lopez, Karla Lopez Aguiniga, Estefany Maria, Jessica E. Rodriguez, Monisha Sebastin, Nehama Teitelman, Dana Watnick, Nicole M. Yelton, Avinash Abhyankar, Noura S. Abul-Husn, Aaron Baum, Laurie J. Bauman, Jules C. Beal, Toby Bloom, Charlotte Cunningham-Rundles, George A. Diaz, Siobhan Dolan, Bart S. Ferket, Vaidehi Jobanputra, Patricia Kovatch, Thomas V. McDonald, Patricia E. McGoldrick, Rosamond Rhodes, Michael L. Rinke, Mimsie Robinson, Arye Rubinstein, Lisa H. Shulman, Christian Stolte, Steven M. Wolf, Elissa Yozawitz, Randi E. Zinberg, John M. Greally, Bruce D. Gelb, Carol R. Horowitz, Melissa P. Wasserstein, Eimear E. Kenny

## Abstract

**Background:** Increasingly, genomics is informing clinical practice, but challenges remain for medical professionals lacking genetics expertise, and in access to and clinical utility of genomic testing for minority and underrepresented populations. The latter is a particularly pernicious problem due to the historical lack of inclusion of racially and ethnically diverse populations in genomic research and genomic medicine. A further challenge is the rapidly changing landscape of genetic tests, and considerations of cost, interpretation and diagnostic yield for emerging modalities like whole genome sequencing.

**Methods:** The NYCKidSeq project is a randomized controlled trial recruiting 1,130 children and young adults predominantly from Harlem and the Bronx with suspected genetic disorders in three disease categories: neurologic, cardiovascular, and immunologic. Two clinical genetic tests will be performed for each participant, either proband, duo or trio whole-genome sequencing (depending on sample availability) and proband targeted gene panels. Clinical utility, cost and diagnostic yield of both testing modalities will be assessed. This study will evaluate the use of a novel, digital platform (GUÍA) to digitize the return of genomic results experience and improve participant understanding for English- and Spanish-speaking families. Surveys will collect data at three study visits; baseline (0 months), results disclosure visit (ROR1, +3 months), and follow up visit (ROR2, +9 months). Outcomes will assess parental understanding of and attitudes towards receiving genomic results for their child and behavioral, psychological and social impact of results. We will also conduct a pilot study to assess a digital tool called GenomeDiver designed to enhance communication between clinicians and genetic testing labs. We will evaluate GenomeDiver’s ability to increase the diagnostic yield compared to standard practices, to improve clinician’s ability to perform targeted reverse phenotyping, and to increase the efficiency of genetic testing lab personnel.

**Discussion:** The NYCKidSeq project will contribute to the innovations and best practices in communicating genomic test results to diverse populations. This work will inform strategies for implementing genomic medicine in health systems serving diverse populations using methods that are clinically useful, technologically savvy, culturally sensitive, and ethically sound.

## BACKGROUND

With the precipitous drop in the cost of genomic sequencing technology over the past two decades, genomic information is increasingly informing clinical decision-making across health systems(1). There are currently over 5,500 single gene disorders and traits with a known molecular etiology (https://www.omim.org/statistics/geneMap). Since 2009, targeted gene panels (TGPs) and exome sequencing (sequencing some or all of the protein-coding regions of the genome, respectively) have been increasingly used for the diagnosis of these individually rare, but collectively common disorders. While the majority of clinical sequencing currently uses panels or exomes, there is an increasing number of pilot programs using more advanced genetic modalities, such as whole genome sequencing (WGS), which has the potential to capture all classes of genetic variation in one analysis(2–4). These advancements in genomic sequencing and testing have sparked innovation in methods for scaling genomic medicine services across health systems.

Although genomic testing is offered to patients in clinical care more routinely, a number of barriers remain to successful implementation of genomic medicine, particularly for racially and ethnically diverse populations. Genomics research in the past has been conducted predominantly in individuals of European ancestry (5,6), which has led to significant disparities in the accuracy and clinical utility of genomic sequencing in non-European populations(7–9). Non-European individuals are more likely to receive variants of uncertain significance or misclassified results, which has been demonstrated in the context of genetic testing for hypertrophic cardiomyopathy and hereditary cancer risks in under-studied populations(10–13). Evidence suggests that this bias is likely to persist in the ongoing and upcoming efforts to sequence entire genomes. Diversifying the data pool in genomics research is a major initiative of federal funding agencies such as the National Human Genome Research Institute (NHGRI) with the goal of enhancing the accuracy, utility, and acceptability of genomic testing in clinical care of diverse populations(14).

Communication of genomic information in health systems poses another significant challenge to genomic medicine implementation(15). For the clinical benefits of genomic testing to be realized, results must be communicated effectively to adults, and families of children, undergoing this testing(16). Genomic test results can be complicated, and vital information can be lost if results are not conveyed in a way that patients and families understand. More studies are emerging to develop and evaluate strategies for communicating genomic results in diverse populations. In New York City (NYC), investigators have evaluated the use of a brief educational program to improve knowledge of complex genomic concepts in a predominantly Hispanic community(16,17). To extend the cultural competence of genetic counseling methods, studies have also designed and assessed the use of culturally tailored genetic counseling methods to convey unique aspects of breast cancer in African-American women(18). Other studies have explored the use of narrative educational tools for at-risk Latinas to improve psychosocial outcomes(19,20). The vast majority of literature on genetic counseling interventions in diverse populations has focused mainly on cancer genetics(21), and there is a dearth of literature on barriers to communication in racially and ethnically diverse pediatric patient populations and their families. Digital applications that display personalized genetic testing results in a way that addresses low-health literacy, includes images, and provides information in both English and Spanish can be leveraged to improve understanding.

A further barrier to broad adoption of genomic medicine is that until recently, this discipline has been the province of a small minority of specialized physicians trained in genomics. The majority of physicians do not receive extensive training in genomics. For example, a survey of United States physicians including generalists and specialists, found that 79% and 69% of primary-care and non-primary-care physicians, respectively, report that “lack of knowledge about genomic medicine” is a barrier to its incorporation in practice(22,23). In a recent study of medical students, 79% felt that it was important to apply genomics to clinical care, but only 6% thought that their medical education had adequately prepared them to practice(24). Genetic testing technologies are moving at a fast pace, and even physicians in a single specialty area can have difficulty keeping up to date on the most effective way to order testing and interpret test reports. For clinical genetic testing laboratories, diagnostic pipelines involve a manual curation step that jointly considers the patients’ genomic variation in the context of the clinical indication for testing and the patient’s phenotypic manifestations, as provided by the physician. However, the input of the clinician’s assessment is usually limited to a few short descriptions on a test requisition that the laboratory personnel have to interpret. Interactions between clinical testing labs and ordering providers must be improved to facilitate interpretation of genomic test results.

The NYCKidSeq study is a genomic implementation research program that will assess strategies for enhanced communication of genomic information in health systems and evaluate the utility of advanced genomic sequencing technology for increased diagnosis. The study will recruit 1,130 children with suspected genetic disorders in historically underserved racially and ethnically diverse communities of NYC. This is a multi-institutional research program with three participating sites; the Icahn School of Medicine at Mount Sinai (MS), Albert Einstein College of Medicine/Montefiore Medical Center (EM), and the New York Genome Center (NYGC). NYCKidSeq has three major aims: 1) to develop and evaluate the effectiveness of a novel digital application, GUÍA, to improve communication of genomic test results to patients in a randomized controlled trial (RCT); 2) to compare the diagnostic yield of WGS versus TGPs in a racially and ethnically diverse cohort; and (3) to evaluate the utility in a pilot study of the novel digital application, GenomeDiver, to enhance the interpretation of sequencing data by laboratory personnel, to direct reverse phenotyping by clinicians, and to enhance the communication between clinical and laboratory personnel in the diagnostic process. The study will also examine the utility of genetic ancestry in clinical diagnostic pipelines, evaluate costs associated with genetic testing, and assess provider attitudes toward genomic medicine. This study is one of six clinical sites funded as part of the Clinical Sequencing Evidence-Generating Research (CSER2) consortium, jointly funded by NHGRI and the National Institute of Minority Health and Health Disparities (NIMHD) (25).

## METHODS/DESIGN

### Study Design Overview

Figure 1 shows the NYCKidSeq project study schema illustrating the flow of enrollment from participant referral to administration of the last parental survey. NYCKidSeq is an RCT evaluating the use of GUÍA to facilitate the return of genomic results compared to standard of care (SOC) genetic counseling. Outcomes to be assessed are parental understanding, satisfaction, feelings about the results, and participants’ subsequent behavior. Surveys will collect data at three study visits: baseline (0 months), results disclosure (ROR1, approximately +3 months), and follow up (ROR2, approximately +9 months). WGS and TGPs will be performed for diagnostic purposes in 1,130 children and young adults with specific, suspected genetic disorders in an effort to assess clinical utility and compare diagnostic yield of both testing methods. Prior to the launch of the RCT, a lead-in pilot phase of 30 participants was conducted to solicit input from families regarding the survey instruments and GUÍA. In designing this study, stakeholders were engaged at key stages of development to facilitate successful implementation of this genomic medicine program and contribute to its cultural appropriateness and sensitivity. As a member-site of the CSER consortium, the funding source has a role in the design of this study with regard to its recruitment goals and outcome measures.

**Figure 1.**
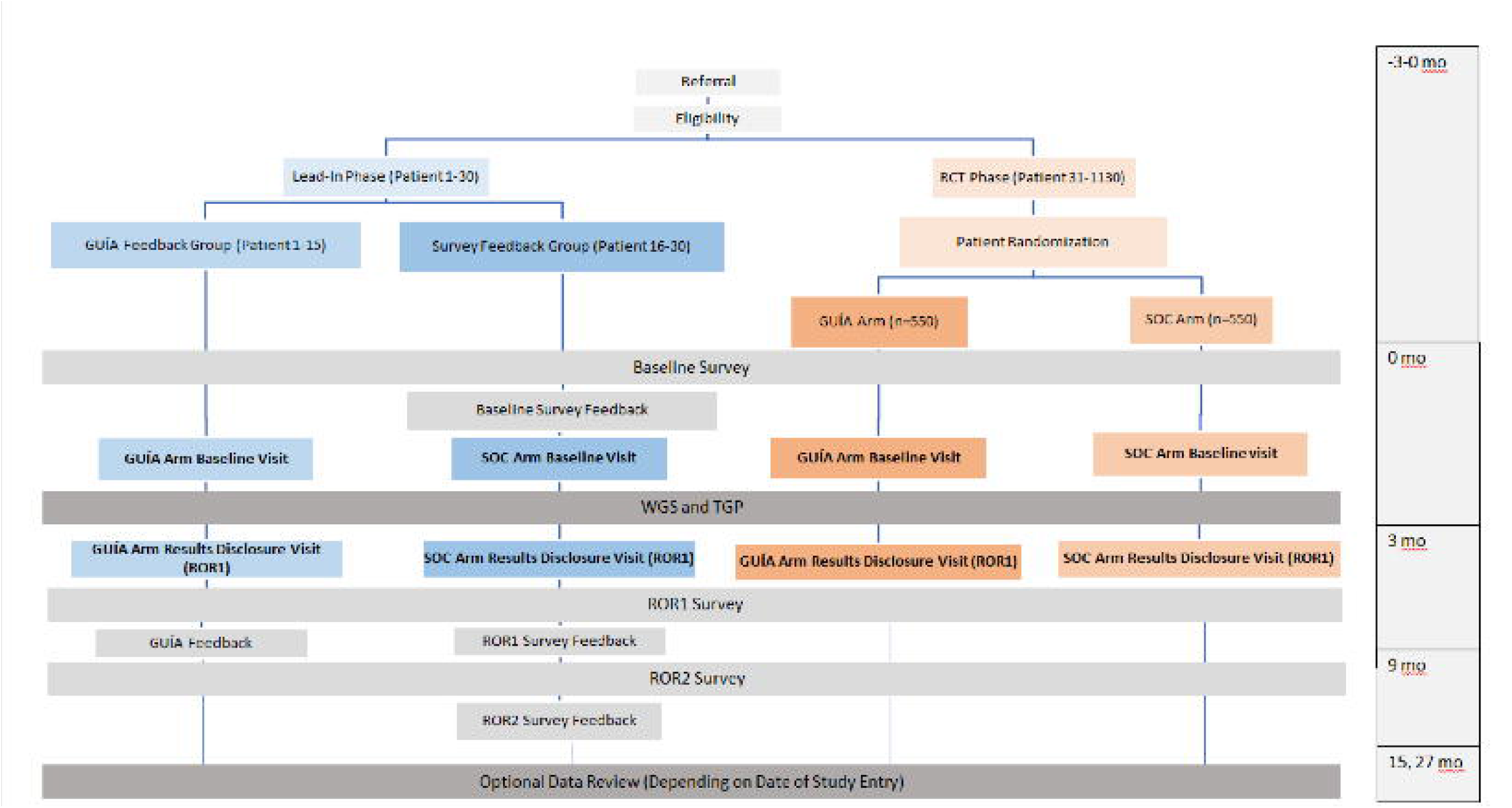
NYCKidSeq study design. The NYCKidSeq study involves two phases, a lead-in phase and a randomized controlled trial (RCT). Participants in both phases receive TGP and WGS testing, and complete surveys at baseline, after result disclosure (ROR1 Survey), and 6-months after result disclosure (ROR2 survey). In the Lead-in Phase, participants (N=30) are enrolled into either the GUÍA feedback arm where participants complete a structured feedback interview after completion of the ROR1 survey or survey feedback arm where participants provide survey feedback after completing each survey. In the RCT phase, participants (N=1100) are randomized to the GUÍA arm or the standard of care arm (SOC). Participants in the GUÍA arm receive results disclosure genetic counselors using GUÍA versus participants in the SOC arm who receive SOC genetic counseling.

## STUDY POPULATION

### Recruitment, Enrollment and Sample Size

Potential participants are receiving medical care under a physician at a participating institution (MS or EM). Participants and their families are introduced to the study by their physician during a routine clinic visit, phone call, or during an in-patient admission. Potential participants who express interest in the study and verbally consent to being contacted by study staff are referred to the study team. Study staff confirms the participant’s eligibility and obtains informed consent to complete the baseline parental survey during an in-person encounter. Surveys are conducted in either English or Spanish, depending on participant preference. Informed consent for WGS and TGP testing is obtained by a genetic counselor during an in-person baseline encounter in the participant’s preferred language (English or Spanish), and assent is obtained from capable children. Enrollment is achieved after a blood draw for the proband’s sample for genetic testing has been collected. At the same time, blood is drawn from biological parents. If one or more biological parents are not available during the visit, a saliva kit is mailed to the home address to collect a saliva sample. The enrollment target is 1,130 participants, including the lead-in phase. Participants enrolled in the RCT receive $80 in gift cards for completing all three study visits and those enrolled in the lead-in phase receive $120 in gift cards. Referring physicians do not receive compensation for their participation.

### Inclusion and Exclusion Criteria

Table 1 presents the inclusion and exclusion criteria for NYCKidSeq participants. All participants are followed by a physician in the participating institutions. Patient participants are 0-21 years of age; young adults (18-21) who are cognitively intact are included in this study provided that their parent(s) or legal guardian(s) also agree to participate. All participants have a currently undiagnosed, suspected genetic cause of their specific neurologic, immunologic, or cardiac disorders. Specifically, participants have at least one of the following: seizure disorder, intellectual disability, global developmental delay, congenital heart disease, cardiomyopathy, cardiac arrhythmia, or features of a primary immune deficiency. Patients will be excluded if they have a known molecular genetic diagnosis, an obvious genetic diagnosis based on clinical features, or if they have undergone a bone-marrow transplant. Inclusion of children of European ancestry is capped at <40% of total participants to ensure that at least 60% of participants are from underserved populations, consistent with the requirements of this funding opportunity.

**Table 1.** Inclusion and exclusion criteria for the NYCKidSeq project.

| **Inclusion Criteria** |
| --- |
| **Demographic criteria** |
| Infants, children, and young adults ($\leq 21$ yo)<br>English- or Spanish-speaking parent or legal guardian<br>Patient at MS or EM<br>Willingness to attend all in-person study visits |
| **Clinical criteria** |
| Currently undiagnosed<br>Suspected genetic cause of a neurologic, immunologic, or cardiac disorder |
| **Prior genetic testing** |
| Results from previous TGP and/or WGS returned $> 3$ months prior to enrollment<br>Results from previous TGP and/or WGS must be negative or identified only a single variant in a gene associated with an autosomal recessive disorder<br>If parents received any genetic counseling for any reason, it must have occurred $> 3$ months prior to enrollment |
| **Exclusion Criteria** |
| Child currently participating in a different genetic sequencing study that includes genetic counseling and/or return of results before the participant's ROR2 visit<br>Known or likely molecular genetic diagnosis for their neurologic, immunologic, or cardiac disorder<br>Bone-marrow transplant |

### Engagement with Diverse Populations

The NYCKidSeq project is recruiting children, young adults and their families from low-income and minority communities which are underrepresented in genomics research and are frequently the last to benefit from advances in research and technology. Participants of all racial and ethnic backgrounds who speak English or Spanish are included with the following distribution of race/ethnicity expected: approximately 1/3 Black/African ancestry; 1/3 Latino/Hispanic ancestry, and 1/3 White/European ancestry.

### Engagement of Non-English-Speaking Patients

Recruitment and retention materials (NYCKidSeq website, brochures, and participant letters), study documents (informed consent documents and surveys), and GUÍA are offered in Spanish and English. Study materials were translated by study staff of Latin American and European descent into Spanish and five Spanish dialects: Mexican, Cuban, Dominican, Puerto Rican, and Castilian. All grew up in exclusively or mostly Spanish-speaking homes, completed Spanish coursework in high school or college, and have worked on research projects that recruited Spanish-speaking participants of a variety of ages, countries of origin, and literacy levels. All had assisted with translation and administration of study materials for prior projects. Staff consulted several online Spanish translation resources such as Word Reference (26) or Linguee (27), as needed for development of multi-dialect compatible content. GUÍA text that is not patient specific was translated by study staff. Participant specific GUÍA text is translated by a study genetic counselor using Google Translate and then reviewed by Spanish-speaking staff to ensure accuracy. Translated survey measures and GUÍA were piloted during the lead-in phase of the study to obtain parents’ feedback on the understandability of the translated text.

### Engagement with Genomic Stakeholder Board

NYCKidSeq engaged the Mount Sinai Genetics and Genomics Stakeholder Board including community leaders of color, clinicians and researchers working together for several years on bridging the gap between academia and communities likely to benefit most from genomic scientific discovery. They participated in designing the study and conceptual framework, consent procedures, patient educational materials, surveys, and recruitment strategies and materials. They meet on a bi-monthly basis to discuss study status, provide feedback to recruitment and retention challenges and assist with analysis of study data, using principles of community-based participatory research to guide their work and ensure meaningful participation(28,29).

### Clinical Genomic Testing

Participants enrolled in NYCKidSeq receive clinical WGS as well as appropriate TGP testing. Proband and biological parental samples, when available, are collected and sent to the NYGC for WGS analysis and Sema4 genetic testing laboratory for TGP analysis. WGS with mean coverage of at least 30x is performed on the NovaSeq platform and is performed as single proband, duo or trio sequencing depending on availability of parental samples. TGP tests offered through the study include neurodevelopmental (448 genes), immunodeficiency (250 genes), and cardiovascular (241 genes) gene panels and are performed as proband sequencing. For probands with symptoms in more than one specified disease area of interest to this study (neurological, cardiac, or immunologic), multiple TGP tests may be ordered. Participants and biological parents may opt-in to receiving secondary findings from WGS testing. For secondary findings, this study is reporting expected pathogenic variants in the 59 genes that the American College of Medical Genetics and Genomics (ACMG) recommends laboratories report (30). Sequencing analysis and variant classification occur at the laboratories using their individual variant interpretation pipelines, and Sanger validation of suspected pathogenic variants is performed. Separate clinical reports are released for each test ordered (i.e., participants will receive at least two test reports). Sema4 and NYGC are Clinical Laboratory Improvement Amendments (CLIA)-certified and approved by the New York State Department of Health to perform TGP and WGS for clinical purposes.

### RCT Intervention (GUÍA)

The Genomic Understanding, Information and Awareness (GUÍA) digital application was developed from formative research as part of the NYCKidSeq study. GUÍA facilitates delivery of individualized genomic results and clinical information to participants and families by allowing genetic counselors to walk participants through their genomic test results in a personalized, highly visual and narrative manner. GUÍA consists of distinct pages with sub-tabs representative of the essential components of a genetic counseling result disclosure session. This includes summaries of the proband’s primary and secondary genomic results, recommendations for next steps for clinical care, inheritance information, educational modules to learn more about the basics of DNA and genomic sequencing, and web links to support groups and related resources. GUÍA presents information in a modular way, allowing the participant to control the depth of the information provided during the session. It can display text in both English and Spanish to increase accessibility for a greater number of participants and their families.

### Pilot Intervention (GenomeDiver)

GenomeDiver is a digital application developed as part of the NYCKidSeq study (31). Using the GenomeDiver web-based platform, the ordering provider is presented with Human Phenotype Ontology (HPO) terms that help to discriminate the candidacy of the highest-ranked DNA sequence variants potentially causing the patient’s phenotype. Following this reverse phenotyping, the enhanced phenotypic information is then used to re-prioritize variants, in turn generating a list of diseases for assessment by the clinician. The additional phenotypic information and any diseases flagged by the clinician as potentially matching the patient’s presentation are then returned to the diagnostic laboratory to inform their interpretation of genomic test results, with the goal of improving genomic diagnostics.

## STUDY ARMS

Participants are randomized to one of two study arms: the control arm was designed to approximate SOC genetic counseling for results disclosure; and the GUÍA arm. “Standard of care” genetic counseling in a research setting has not been well defined in the literature. It is challenging to define SOC in genetic counseling as genetic counselors practice in a variety of clinical settings, both in-person and remotely. For the purposes of this study, SOC genetic counseling for results disclosure consists of contracting, review of the purpose of the genomic testing, and disclosure of the child’s genomic test results. For positive test results, the genetic counselor describes the diagnosis, associated symptoms, management recommendations, and life expectancy, if known. The genetic counselor then discusses the inheritance pattern, recurrence risks, and identifies at-risk family members who may also require/consider testing. In the case of negative results, the genetic counselor discusses the implications of such a result, such as the possibility that there is a genetic cause for the child’s symptoms that was unable to be identified at this time by this testing. For ambiguous results, the genetic counselor explains the meaning and uncertainty associated with these types of results and provides recommendations for continued disease management. The genetic counselor also discloses any secondary findings to participants who opted to receive those results. Throughout the session, explanations are accompanied by visual aids at the discretion of the genetic counselor. Psychosocial concerns are addressed throughout the encounter. The genetic counselor provides medical and support referrals, when appropriate. As the WGS and TGP are approved for clinical purposes, reports are given to the families and incorporated into their medical records, and shared with referring physicians. Letters or handouts that summarize the sessions are provided after disclosure of results to explain the findings to the patient, family, physicians and/or insurance companies for additional services. Patient or family support resources such as syndrome or symptom specific parent support groups, research and awareness organization websites, and/or publicly available information booklets are provided based on the needs of the family. Families are encouraged to return to their referring provider for continued post-test clinical care.

During the GUÍA genetic counseling results disclosure, the genetic counselor follows the same procedures as those outlined for the SOC arm using GUÍA during the genetic counseling session to facilitate this discussion. Prior to the results disclosure session, genetic counselors personalize GUÍA by inputting genomic test result information, clinical details, inheritance, family implications, medical recommendation, and support referrals. At the close of the results disclosure appointment, the genetic counselor provides the family a hard copy of their child’s personalized GUÍA report.

## PROCEDURES

Figure 1 shows the study flow and data collection points of the NYCKidSeq project. This includes a lead-in phase of the first 30 participants and the subsequent RCT. The first 30 families who met eligibility requirements and agreed to participate were entered into the lead-in phase of the study. All other participants are enrolled into the RCT and randomized to either the SOC or the GUÍA study arm.

### Lead-In Phase

The participants of the lead-in phase (N=30) were not randomized to a study arm. Instead, they were asked to provide feedback on the surveys or on GUÍA. Participants in the lead-in phase completed the same series of study visits as those in the RCT phase (study visits are described in detail below).

The first 15 participants received genomic results with genetic counseling using GUÍA. After their results session, a trained study team member collected participant feedback on GUÍA using a brief, structured feedback guide to explore parents’ reactions to GUÍA. Feedback was used to address and clarify wording/phrasing, use of images, order of information, amount and detail of information, pace, and potential Spanish translation issues.

The next 15 participants provided input about the surveys. After each study survey was administered, a trained member of study staff gathered participant feedback on the survey using a “think aloud” format. Participant feedback focused on survey question clarity, flow, and order. These participants received genomic results using SOC genetic counseling.

### Randomized Controlled Trial

Participants in the RCT (N=1100) are randomized using a stratified randomization scheme by disease category (cardiac, neurologic, immunologic) and clinical site as seen in Figure 2. Randomization occurs prior to the baseline visit (BL) via a randomization module in REDCap applied by a study staff member. The REDCap randomization allocation is not revealed to study staff at any point in the study.

**Figure 2.**
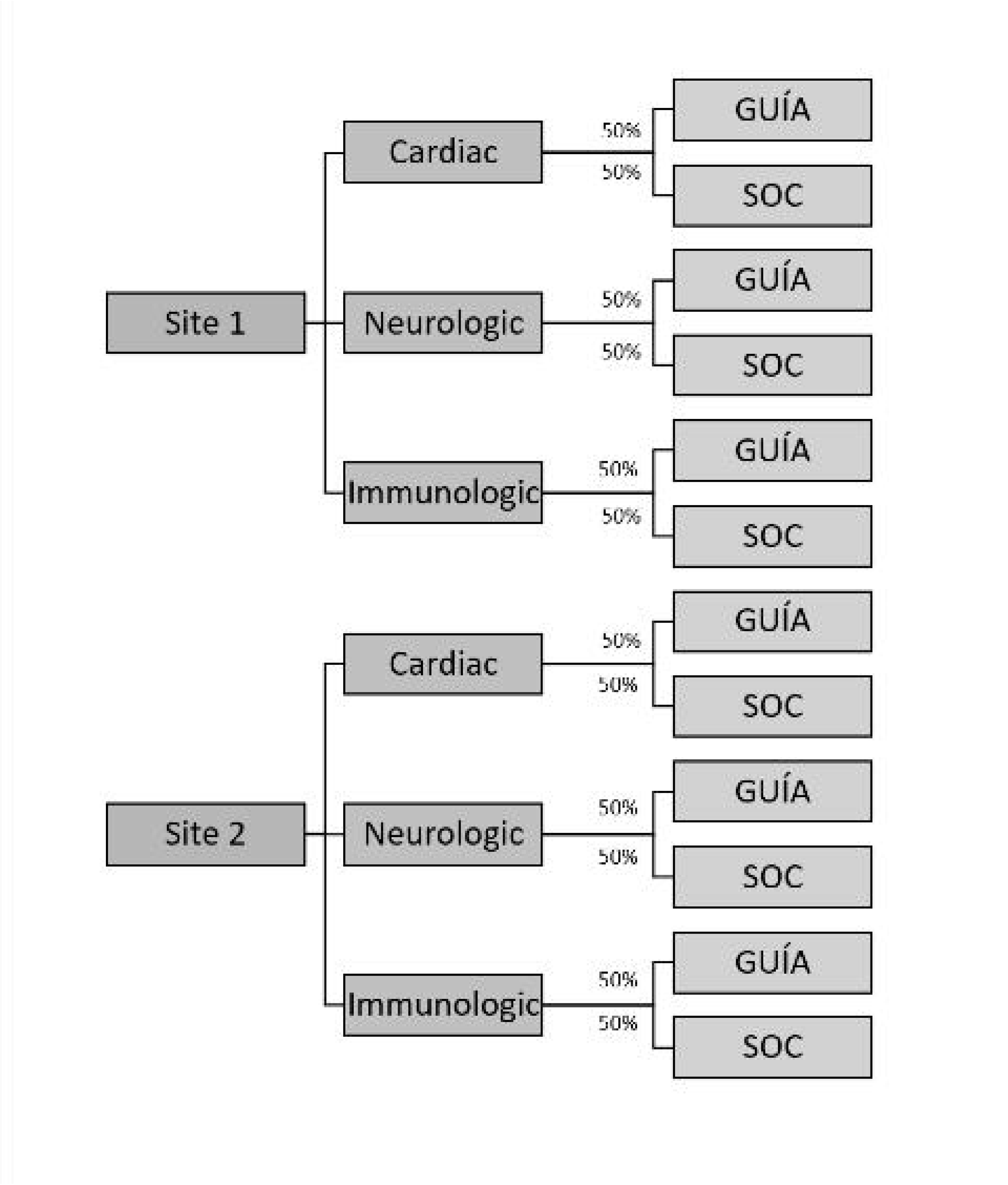
Randomization Schema.

**Figure 3.**
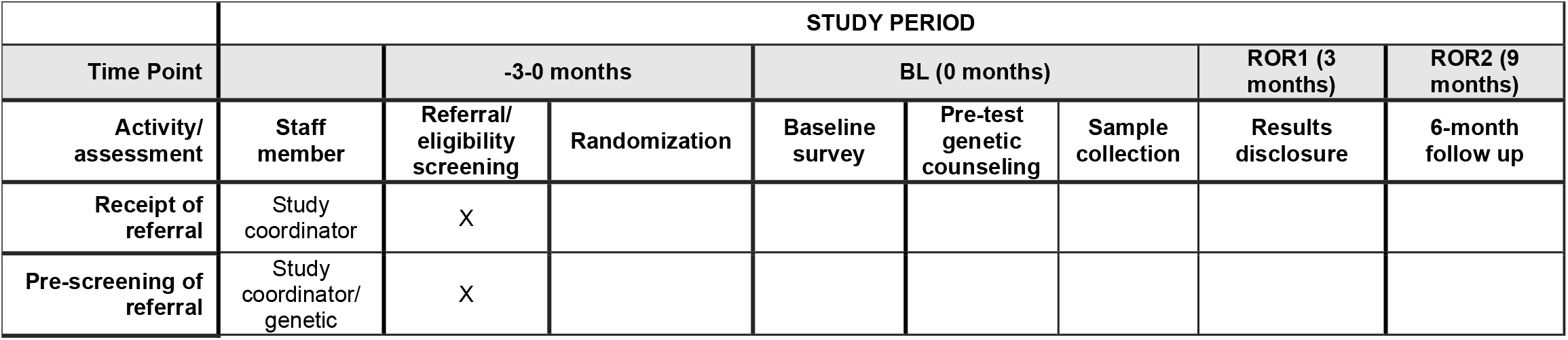
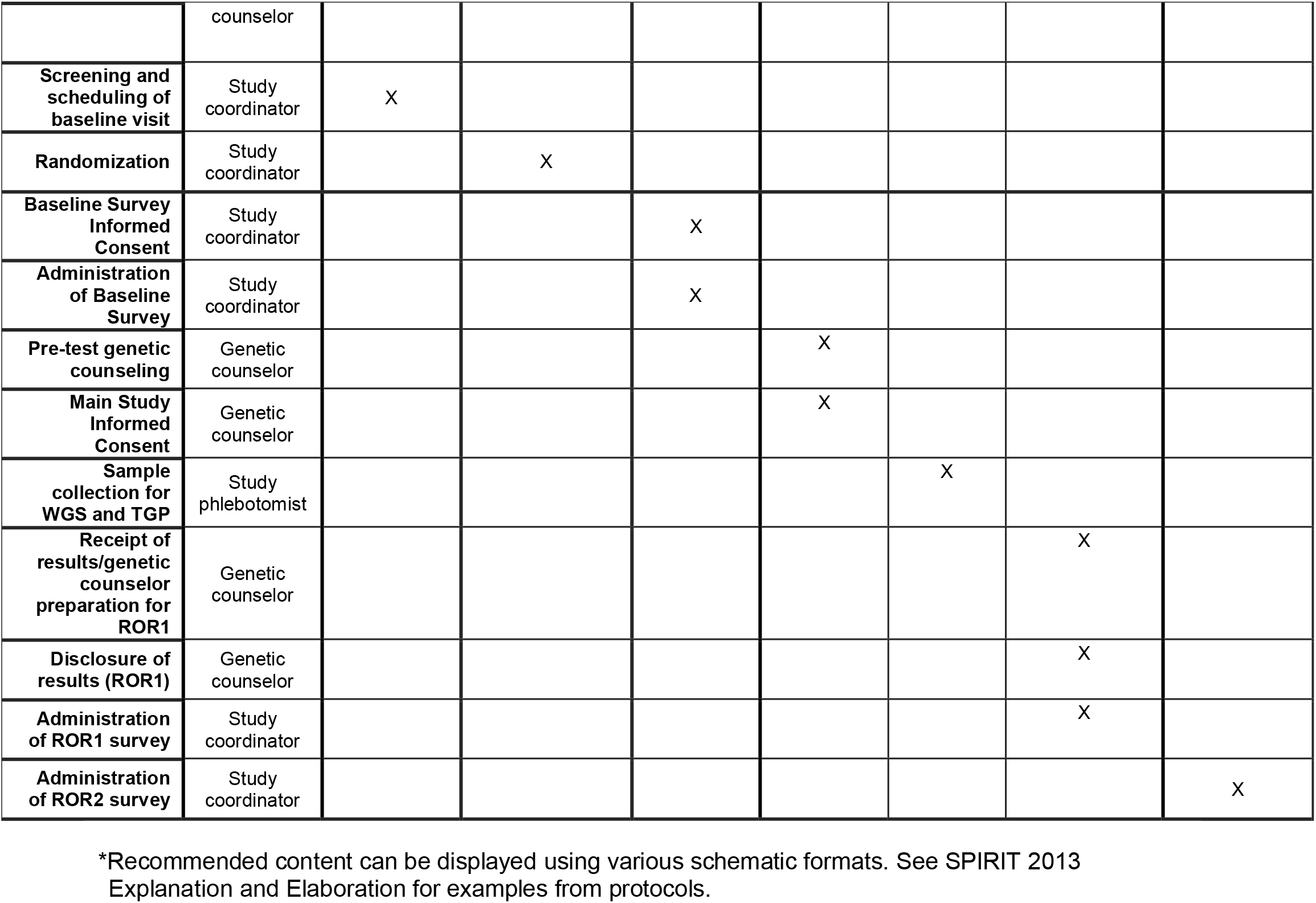
Schedule of forms and procedures (adapted from original SPIRIT table).*

At the BL visit participants complete the baseline surveys with a study staff member and receive pre-test genetic counseling by a genetic counselor designated to their arm. The consenting process consists of the family being educated about the study, the risks, benefits, and limitations of genomic testing, purpose of genomic testing, possible results of genomic testing including the option to receive or decline secondary findings, and potential implications for other family members. As part of the pre-test genetic counseling, the genetic counselor obtains a medical and family history. Families who consent for testing undergo blood or saliva collection for both TGP and WGS. Parents and cognitively intact young adults will also provide permission to use of de-identified samples in future research; sharing of de-identified data in secure, public research databases; and to be contacted by trial investigators for further informational and consent-related purposes. Participants consenting to take part in this study voluntarily agree to indefinite storage of their and their child’s blood and sequencing information by the research study, including NYCKidSeq research teams at Sema4, NYGC, EM, and MS. Samples may be used for either research of clinical purposes if additional testing is needed. Participant can decide to withdraw consent for storage of their or their child’s biological samples at any time by contact the principal investigator. Sample(s) or portions thereof that have not already been used will be destroyed; however, the parent or child’s sample may have already been distributed to other researchers within NYCKidSeq before the request to destroy was received and may not be able to be retrieved.

Once a participant’s results are reported, generally after three months, the results are sent to the genetic counselor. Results are reviewed and shared with the referring physician and/or a geneticist who then shares their interpretation about the significance of the genomic findings as well as their medical recommendations with the genetic counselor. An *ad hoc* discrepancy committee is available to review cases at the discretion of the genetic counselor for cases that have discrepant or unsatisfying results. The committee consists of NYCKidSeq medical geneticists, genetic counselors, laboratory directors, and referring providers. The decision of the discrepancy committee is used as a final diagnostic determination.

Each participant has a one-on-one results disclosure visit with a genetic counselor (ROR1). The referring physician can also participate in the appointment at their discretion. At the results disclosure appointment, the participant is informed of the results and their referring provider’s medical recommendations. The method of genetic counseling delivery depends on the study arm the participant is randomized to. After genetic counseling, participants immediately complete the ROR1 survey with a study staff member. Six months after the ROR1 visit, approximately 9 months after study entry, a follow up visit (ROR2) occurs either by phone or video. During this interaction a study staff member administers the final survey (ROR2 survey).

Each subject’s genetic results may be reviewed every twelve months for the duration of the study. This is because information about genetic variants can change over time, as can the patient’s phenotype. As both types of information contribute to making a diagnosis, a re-analysis that recognizes reclassification of DNA sequence variants in a patient and their current phenotypic presentation can combine to change their original diagnosis. Variant reclassification information is derived by the laboratory from public databases such as ClinVar, while the refined phenotypic information is prompted by and entered into GenomeDiver following a review of electronic medical records. If results are reinterpreted, a new visit is arranged to inform the family of the finding. The visit to review the results is performed by a genetic counselor.

### Pilot Study

We will also recruit 20 referring providers, 6 genetic counselors, and 7 laboratory staff for our GenomeDiver pilot study. Two different interventions will be performed. One is retrospective and facilitates updating participants’ phenotypic information 12 months after ROR1 to help with the review of the original diagnostic laboratory report. The second intervention is part of the ongoing recruitment of patients and occurs as part of the primary analysis of the patient’s genomic sequence. We randomize these patients into two arms, one with SOC, the other with the addition of a GenomeDiver intervention. Following the generation of the annotated Variant Call Format (VCF) file, the clinicians (referring provider and genetic counselor assigned to the patient) are contacted and requested to perform a GenomeDiver session. HPO terms that help to discriminate the variants with the highest Exomiser combined scores (32) are presented to the clinicians and categorized at present, absent or uncertain for that patient. Potential diseases present in the patient are then displayed for clinician evaluation and possible flagging, and the enhanced, updated information is then returned to the diagnostic laboratory.

## STUDY OUTCOMES

### RCT Survey Measures

The NYCKidSeq project is assessing participant outcomes through surveys administered at BL, prior to pre-test counseling and consent for the genomic testing; following the results disclosure genetic counseling during ROR1; and 6-months after disclosure of results at ROR2. The primary study outcome is the participant’s perceived understanding of genomic testing results, with comparison of results in SOC arm to GUÍA arm. The secondary study outcomes are objective understanding of genomic testing results and understanding of medical follow up, the actionability of genomic results, and adherence to medical follow up recommendations, with comparison of results in SOC arm to GUÍA arm. Additional participant outcomes focus on six domains: (a) attitudes towards genomic testing, (b) perceived utility, (c) psychological, (d) behavioral, (e) social, (f) economic impact of genomic testing. With the exception of economic impact, all outcomes will be compared between the two study arms. The CSER consortium harmonized survey measures so that CSER projects, when possible, administer standardized survey measures (25) to facilitate combining data into a single data set for cross-consortium analysis. Table 2 summarizes participant outcomes being assessed across the three time points. When possible, previously validated measures were used. The BL and ROR1 surveys are administered in-person by a study staff member while the ROR2 survey is administered by a study staff member over the phone or video. Surveys are administered in either English or Spanish. All survey response data is entered and maintained in the REDCap database.

**Table 2.** NYCKidSeq participant outcomes by survey timepoint.

| **VARIABLE** | **SOURCE** | **BL<sup>1</sup>** | **ROR1<sup>2</sup>** | **ROR2<sup>3</sup>** |
| --- | --- | --- | --- | --- |
| **Primary Outcome** |  |  |  |  |
| *Perceived understanding of genomic testing results* | NYCKidSeq developed measure (novel) | - | X | X |
| **Secondary Outcomes** |  |  |  |  |
| *Objective understanding of genomic testing results* | NYCKidSeq developed measure (novel) | - | X | X |
| *Medical actions and non-medical/patient-initiated actions attributable to genomic testing* | CSER developed measures (novel): Attributable to Genomic Testing (RMA) and Patient-Initiated Actions Attributable to Genomic Testing (PIA) | - | - | X |
| **Attitudes** |  |  |  |  |
| *Satisfaction with mode of delivery* | CSER developed measure (novel) adapted from Patient Assessment of cancer Communication Experiences (PACE) (33,34) | - | X | - |
| *Satisfaction with results* | Satisfaction with information about medicine (SIMS) (35) | - | X | - |
| *Attitudes toward genetic testing* | Adapted from Genetic testing to Understand and Address Renal Disease Disparities (GUARDD) study (36,37) | X | X | X |
| *Empowerment* | Adapted from GUARDD study (36) | X | X | X |
| *Decisional conflict* | Decisional Conflict Scale (Low Literacy) (38) | X | X | X |
| **Perceived Utility** |  |  |  |  |
| *Impact of genomic testing on health status* | Functional status II-R (child) (39) | X | - | X |
| *Impact of genomic testing on quality of life* | Child Health Utility Instrument (CHU9D; parent as proxy) (40); SF-12 health survey (for parent) (41) | X | - | X |
| *Clinical utility* | Patient Reported Utility (PrU) (42) | - | X | X |
| **Psychological Impact** |  |  |  |  |
| *Feelings about genomic testing results* | Feelings About Genomic Testing Results (FACToR) (43) | - | X | X |
| *Uncertainty* | Perceptions of Uncertainties in Genomic Sequencing (PUGS) (44); FACToR subscale (43) | - | X | X |
| *Depression* | 8-item Patient Health Questionnaire depression scale (PHQ-8) (45) | X | X | X |
| *Anxiety* | Generalized Anxiety Disorder Screener (GAD-2) (46,47) | X | X | X |
| *Perceived stress* | Perceived Stress Scale 4-item (PSS-4) (48) | X | X | X |
| *Self-Efficacy* | Decision Self-Efficacy Scale (49) | X | - | - |
| *Patient activation* | Short Form Patient Activation Measure (PAM) (50) | X | - | - |
| *Decisional regret* | Decision Regret Scale (51) | - | X | X |
| **Behavioral Impact** |  |  |  |  |
| *Information seeking* | CSER developed measure (novel); Adapted from Psychological Adaptation to Genetic Information Scale (52) | - | X | X |
| *Family communication* | CSER developed measure (novel) | - | - | X |
| **Social Impact** |  |  |  |  |
| *Support* | Low-Literacy Decisional Conflict Scale (Q6 and Q8) (53) | X | X | X |
| *Access to care* | CSER developed measure (novel) | X | X | X |
| *Life chaos* | Chaos Scale (54) | X | - | - |
| *Family & community* | Medical Outcomes Study Social Support Survey (mMOS-SS) (55) | X | - | - |
| *Quality of life ascertainment (for child)* | PedsQL Parent Proxy Generic Core (56); EuroQol-Visual Analog Scale (VAS) (57) | X | - | X |
| **Economic Impact** |  |  |  |  |
| *Cost/Value* | CSER developed measure (novel) | - | X | X |
| *Healthcare utilization* | Self-reported Utilization of Health Care Services (58) | - | X | X |
| **Sociodemographic Factors** |  |  |  |  |
| *Literacy; Numeracy* | BRIEF Health Literacy Survey (59); Subjective Numeracy Scale (SNS-3) (60) | X | - | - |
| *History of receiving genetic testing* | Adapted from the GUARDD study (36) | X | - | - |
| *Trust in healthcare system* | CSER developed measure (novel) adapted from Health Care System Distrust Scale (61) | X | - | - |
| *Health beliefs* | Brief Illness Perception Questionnaire (IPQ) (62) | X | - | - |
| *Child and Parent: sex, age, race/ethnicity, country of origin, language, insurance status, residential history, zip code* | CSER developed measure (novel); Adapted from HCHS/SOL Personal Information Questionnaire (63) | X | - | - |
| *Parent only: Education level, employment, income, household, marital status* | CSER developed measure (novel); Adapted from HCHS/SOL Personal Information Questionnaire (63) | X | - | - |
| *Grandparents of child: Residential history* | Adapted from HCHS/SOL Personal Information Questionnaire (63) | X | - |  |

<sup>1</sup>BL = Baseline survey
<sup>2</sup>ROR1 = Return of results, visit 1 survey
<sup>3</sup>ROR2 = Return of results, visit 2 survey

### Pilot Study measures

The quantitative outcome sought from the GenomeDiver interventions is whether it led to an increase in the rate at which the genetic test yields a clinical diagnosis compared with SOC. We are also testing other outcomes. Laboratory personnel will be asked to assess how long analyses took for individual patients, how many variants were considered per patient, whether they gained insights into the ability of clinical colleagues to identify specific phenotypes, and whether the prospective GenomeDiver intervention overall changed the time to issue a report, as a concern is that introducing a delay in analyzing the VCF file while waiting for GenomeDiver input could lead to the report being overdue. Clinicians will be asked whether the HPO terms appeared to be appropriate for the presentation using yes, no, or maybe designations, the time spent performing sessions, the diagnostic value and ease of interpretation of HPO terms, whether any further testing was prompted by the suggested HPO terms, and any difficulty categorizing specific HPO terms because of the race/ethnicity of the patient. The analysis will also include testing whether the referring provider’s specialty or with patient properties, such as age, number of notes in electronic medical record or length of time in the health care system are associated with the HPO term categorization patterns.

### Diagnostic Yield and Comparison of WGS to TGP

The overall diagnostic yield of the genomic testing will be calculated as the percentage of NYCKidSeq participants with definitive or likely positive diagnoses. Individual diagnostic yield will be calculated for WGS and TGP tests as well as by disease category (neurology, cardiology, immunology). We will also investigate the concordance between the two testing modalities. Lastly, we will assess the diagnostic yield of both tests among race/ethnic groups. Genomic testing result categorization for both testing modalities is maintained in the REDCap database.

## CONFIDENTIALITY

Genetic counselors and other study clinicians access the participants’ medical record to obtain relevant clinical information, such as medical diagnoses and previous genetic test results. This information is reviewed and collected in accordance with The Health Insurance Portability and Accountability Act (HIPAA). The following procedures are used at MS and EM safeguard data: 1) train staff on data sensitivity and safeguards; 2) store and process sensitive hard copy in a centralized location; 3) secure sensitive hard copy in locked files when not in use; 4) remove names, addresses, and other direct identifiers from hard copy and computer-readable data if they are not necessary for participant tracking; 5) destroy all identifiable links to data after accuracy has been verified and final analyses have been completed; and 6) protect the patient information file, secured in our file server, by Microsoft NT encrypted password and a separate password to access the database file on the server.

Limited identifying information of consented participants is stored in a web-based REDCap database. The REDCap server is managed by Mount Sinai IT and is firewall protected. User access to the database for study personnel is managed by the study project manager. Data access for study personnel is limited to their site participants and what is required for their roles on the project. The NYCKidSeq program gives participants the option to consent to share de-identified genetic and related clinical information to be shared with other CSER investigators and access-restricted scientific databases.

As this study involves genetic testing for diagnostic purposes, WGS and TGP results are entered into the medical record along with the accompanying genetic counseling chart notes. These documents are maintained in the participant’s permanent medical record. The remaining clinical research records including IRB documentation are retained for at least three years at MS and at least 25 years at EM after the clinical research study is completed, consistent with NIH and FDA policies, or longer if required by Mount Sinai. Upon completion of this period, documents will be shredded and disposed of in accordance with hospital requirements.

### Analysis of Outcomes

#### Lead-in Phase

Data from the feedback sessions of parents performed during the lead-in phase of the study (N=30) to learn about the GUÍA (N=15) and to identify any issues with the surveys (N=15) will be reviewed. Any useful feedback from these sessions will be incorporated into GUÍA and the surveys.

#### RCT

Descriptive statistics will be calculated for quantitative survey instruments in the baseline, ROR1 and ROR2 surveys. In the case of missing data, when survey measures contain summary scores, a mean score will be calculated based on the responses provided. We will adjust for covariates, including age, sex, and race/ethnicity where appropriate. Repeated measures of analysis of variance (ANOVAs), chi-squared test or regression models will be fit to the data in a simple paired design (N=550 on each arm) to assess and identify significant improvements in parental understanding, satisfaction, and feelings about the results, and their subsequent behavior in the SOC group compared to GUÍA group. A statistical significance criterion of p< 0.05 (after adjustment for multiple testing) will be used for all analyses.

#### Diagnostic yield

We will also perform analysis to compare the clinical utility and diagnostic yield of WGS compared to TGP by comparing the results status (positive, negative, and uncertain) via each modality. We will focus our analysis on concordance, accuracy and reproducibility as being most important for clinical utility. We will also examine differences in diagnostic yield of pathogenic, likely pathogenic or uncertain variants across race/ethnicity groups.

## DISCUSSION

The NYCKidSeq study aims to recruit 1,130 children and their families, predominantly from Harlem and the Bronx areas of NYC to a RCT. Recruitment began January 31, 2019 and is expected to be completed by May 31, 2021. Eligible children are suspected of having an undiagnosed genetic disorder in three disease categories: neurologic, cardiovascular, and immunologic. Two clinical genetic tests will be performed for each participant, either proband, duo or trio WGS (depending on sample availability) and proband TGPs. Clinical utility, cost and diagnostic yield of both testing modalities will be assessed and compared. This study will evaluate the use of a novel, digital platform called GUÍA to digitize and standardize the return of genomic results and improve participant understanding, designed for both English- and Spanish-speaking families. The outcomes are parental understanding of and attitudes towards receiving genomic test results for their child, and behavioral, psychological and social impact of genomic results. We will also conduct a pilot study to assess a digital tool called GenomeDiver designed to enhance communication between medical professionals and genetic testing labs, and evaluate its ability to increase diagnostic yield when used as a means to improve communication to share phenotypic and genotypic information, compared to standard practices.

There are several limitations to the study design, which includes the lack of blinding in the RCT. Participants and study staff are aware of participants’ randomization status as participants in the intervention arm are asked about their experiences using GUÍA specifically. In addition, GUÍA is unavailable in languages other than English and Spanish. After evaluating the use of GUÍA in these languages, we hope to expand the usability of GUÍA in other languages to be more reflective of the immense linguistic diversity of NYC.

In summary, the NYCKidSeq Study is investigating the effectiveness of integrating WGS into the clinical care of diverse and medically underserved children and their families in a variety of healthcare settings and disease specialties. This work is contributing to the broader NHGRI-funded CSER consortium, now in its second funding cycle. Goals of the consortium include assessing the clinical utility of WGS, exploring medical follow up and patient outcomes, and providing new technology to enhance communication of genomic information within health systems and communities, and evaluating patient-provider-laboratory level interactions that influence the use of this technology. The findings from this study, and the broader CSER consortium, will inform a clearer understanding of the opportunities and barriers of providing genomic medicine in diverse populations and clinical settings, and contribute evidence toward developing best practices for the delivery of clinically useful and cost-effective genomic sequencing in diverse healthcare settings.

## Data Availability

De-identified data for this study will be shared in secure, access-restricted scientific research databases called The Database of Genes and Phenotypes (dbGAP) at the National Institutes of Health. Interpretations of clinical significance of variants from genetic testing will be submitted to the ClinVar database at the National Institutes of Health.

## Trial status

NYCKidSeq Protocol version 10, October 15, 2019. Recruitment began January 31, 2019 and is expected to be completed by May 31, 2021.

## List of abbreviations

TGP: targeted gene panel
WGS: whole genome sequencing
GUÍA: Genomic Understanding, Information and Awareness application
SOC: standard of care
ROR: return of results
MS: Icahn School of Medicine at Mount Sinai
EM: Albert Einstein College of Medicine/Montefiore Medical Center
NYGC: New York Genome Center

## Declarations

### Ethics approval and consent to participate

The Institutional Review Boards of Icahn School of Medicine at Mount Sinai and Albert Einstein College of Medicine approved this study protocol. Written, informed consent to participate will be obtained from all participants.

### Consent for publication

A model consent form will be made available upon request.

### Authors’ contributions

EEK, MPW, CRH, BDG, JMG, GAD, CCR, TB, LB, AB, RR, MLR, MR, AR, and SD made substantial contributions to the conception of the study and the design of the work. JAO, KMG, SAS, KED, MAR, NRK, GB, CB, KB, LF, JL, KL, EM JR, MS, NR, DW, NY, AA, NSAH, AB, LJB, JCB, TB, GAD, SD, BSF, VJ, MR, CS, REZ, JMG, BDG, CRH, MPW, and EEK helped in the acquisition, analysis and interpretation of data. JAO, KMG, SAS, NRK, GB, TB, PK, CS, JMG, and EEK contributed to the creation of new software used in the work. JAO, KMG, SAS, KED, MAR, NRK, GB, CB, KB, LF, JL, KL, EM, JR, MS, NT, DW, NY, AA, NSBH, AB, LJB, JCB, TB, CCR, GAD, SD, BSF, VJ, PK, TVM, PMG, RR, MLR, MR, AR, LS, CS, SW, EY, REZ, JMG, BDG, CRH, MPW, and EEK drafted this work or substantially revised it.

## Acknowledgements

The authors would like to thank the following individuals; Hernis de la Cruz, Mimsie Robinson, Kojo Davis, Trinisha Williams, Irma Laguerre, Melvin Gertner, Lynne Richardson, Reymundo Lozano, Wilson Heredia Nunez, David Kaufman, Jessica Oh, Julie Flom, Jao Pedro Matias Lopes, Nicole Ramsey, Maite La Vega-Talbott, Soledad Puente-Guzman, Jules Beal, Paul Levy, Sheri Escalante, Puja Patel, Robert Marion, Zachary Hena, Bradley Clark, Daphne Hsu, Neha Bhatia, Christine Walsh, Karen Ballaban-Gil, Kimberly Reidy, Daniel Lax, Manoj Gupta, Mohamad Saifeddine, Rana Jehle, Tatyana Gavrilova, Gregory Gutin, Nagma Dalvi, Elisa Muniz, Koshi Cherian, Mana Mann, Sarah Van Dine, Maris Rosenberg, Maria Valicenti-Mcdermott, Rosa Seijo, Lisa Shulman, Elisa Muniz, Meghan Keenan Breheney, Michael Satzer, Jennifer Yoffe, Dona Rani Kathirithamby, Sindhu Gupta, Gina Cassel, Parmpreet Kaur, Rachel Eisenberg, Margo Breilyn, Emily Jackness, Arjola Cosper, Joanna Grater, Gina Cassel, Emily Jackness, Isaac Molinero, Jenny Shliozberg Richard Sidlow, Larry Bernstein, Zoya Treyster, Leslie Delfiner, Susan Duberstein, and Farah Dosani. We would also like to thank the parents and children participating in this study.

## Authorship Guidelines

Principal investigators will review topics suggested for presentation or publication. Lead author(s) will be determined by principal investigators. Any disputes regarding authorship will be resolved by the principal investigators.

## Protocol Amendments

Any significant changes to the protocol outlined above must be written in a formal amendment. The amendment must be approved by both IRBs at MS and EM.

## Trial Registration

ClinicalTrials.gov, Identifier: NCT03738098. Registered on November 13, 2018, https://clinicaltrials.gov/ct2/show/NCT03738098

## Trial Sponsor

Icahn School of Medicine at Mount Sinai

## Contact Name

Eimear Kenny, PhD (Principal Investigator)

## Address

Icahn School of Medicine at Mount Sinai, One Gustave L. Levy Pl., Box 1003, New York, NY 10029

